# GLP-1 Receptor Agonist Prescription Patterns in the *All of Us* Research Program

**DOI:** 10.64898/2026.01.13.26344022

**Authors:** Angela Gasdaska, Benjamin Tyndall, Ed Preble, M. Daniel Brannock, Melissa McPheeters, Laura Marcial, Ariba Huda, Josephine M. Egan, Tamara R. Litwin, Lorenzo Leggio, Mehdi Farokhnia, Chandan Sastry, Jennifer Adjemian

## Abstract

**Importance:** Glucagon-like peptide-1 receptor agonists (GLP-1RAs) are fast-growing treatments for type 2 diabetes, obesity, and sleep apnea and are under investigation as potential treatments for many other conditions. The National Institutes of Health’s (NIH’s) *All of Us* Research Program offers a robust observational data source for studying questions related to GLP-1RA use in real-world settings.

**Objective:** This article describes key characteristics of *All of* Us participants who have been prescribed GLP-1RAs. The goals are to present the utility of the *All of Us* data and describe the strengths and limitations of using this resource for future research on GLP-1RAs.

**Design:** Using the *All of Us* Controlled Tier Curated Data Repository version 8 (CDRv8), we provide a descriptive analysis of the cohort with GLP-1RA records using cross-sectional surveys, longitudinal electronic health record (EHR) data, and longitudinal Fitbit data.

**Setting:** The *All of Us* Research Program is a large, federally funded, longitudinal cohort study established in 2018 by NIH. Recruitment efforts are nationwide and target a range of populations to advance precision medicine for all.

**Participants:** Participants are U.S. residents, aged 18 or older at the time of study consent, who were enrolled between May 6, 2018, and October 1, 2023.

**Exposures:** The GLP-1RA cohort included participants with at least two GLP-1RA prescription records on different days at any time point based on their EHRs.

**Main Outcomes:** Frequencies and medians for a range of sociodemographic characteristics, health care utilization patterns, comorbid conditions, GLP-1RA prescription trends, laboratory and observation availability, and Fitbit data.

**Results:** The *All of Us* GLP-1RA cohort is large (n=15 477), with high data availability across a range of relevant data types. These participants are older and have more comorbid conditions than the entire CDRv8 population. Prescription trends indicate rapid uptake of GLP-1RA drugs since 2014.

**Conclusions and Relevance:** *All of Us* CDRv8 is a valuable resource for research on GLP-1RAs across a large, heterogeneous cohort of participants. The variety and availability of data offer many possibilities for future observational, real-world research to address unanswered questions about GLP-1RA use and replicate recent findings generated from other datasets.

**Key points:** *Question:* What data are available and what are the patterns of GLP-1 receptor agonist (GLP-1RA) prescriptions among participants in the *All of Us* Research Program?

*Findings:* In this descriptive cohort study of 633 534 *All of Us* participants, 15 477 participants had at least two records of GLP-1RA prescriptions. These participants tended to be older, have more comorbid conditions, and have higher health care utilization than the *All of Us* population as a whole.

*Meaning:* The *All of Us* Research Program has a robust array of data to support observational studies of people receiving GLP-1RA prescriptions.

## Introduction

Glucagon-like peptide-1 (GLP-1), an incretin hormone that regulates glucose homeostasis, appetite, and food intake, acts as a neurotransmitter involved in brain mechanisms related to satiety, reward processing, stress response, cognition, and neuroinflammation. Several GLP-1 receptor agonists (GLP-1RAs) are approved to manage type 2 diabetes, and some are approved for obesity and to treat related obstructive sleep apnea. GLP-1RAs are receiving considerable attention due to their versatile therapeutic potential across various chronic diseases. Research is ongoing to investigate GLP-1RAs’ impact on cardiovascular, liver, and neurodegenerative diseases; mental health conditions; and substance use disorders, among others. Development of new GLP-1RAs is focused on higher potency and longer duration of effects, fewer side effects, easier administration and improved adherence, and new therapeutic effects.^1^ GLP-1RA prescriptions have grown tremendously in the last decade, with some studies estimating nearly 20% of adults with type 2 diabetes had prescriptions in 2022, up from less than 5% in 2016.^2^ Multiple new GLP-1RAs are under development, including poly-agonists; the number of clinical conditions for which they are being studied continues to expand.^1^

The *All of Us* Research Program is a large, federally funded, longitudinal cohort study established in 2018 by the National Institutes of Health (NIH). Data collected through *All of Us* are from electronic health records (EHRs), surveys, physical measurements, genomic testing, and wearable sensors.^3,4^ Participation is open to all adults in the United States, with recruitment efforts additionally targeting heterogeneous populations to advance precision medicine for all; over 84% of participants are members of communities with historical gaps in research experience, such as populations with unique life experiences and health needs (e.g., older adults, rural populations, individuals with less access to health care). Data are available to approved researchers through the *All of Us* Researcher Workbench, a secure platform for data analysis.

The current release of *All of Us* data is the Curated Data Repository version 8 (CDRv8), which was released on the workbench in February 2025. CDRv8 includes survey responses from over 633,000 participants; physical measurements for over 509,000; genotyping arrays for over 447,000; EHR data from over 393,000; and Fitbit data for over 59,000. Data span from May 2018 through September 2023, although EHR and Fitbit data include earlier records, some from as far back as 1980.^5^

Significant increases in GLP-1RA use during the years of *All of Us* data collection, combined with the breadth of data available, present new research possibilities. Given the size and scope of *All of Us*, these data enable researchers to address important questions related to GLP-1RAs (e.g., duration of use, long-term outcomes, emerging side effects, morbidity) and to test findings generated from other datasets. Of note, genomic data, survey responses, and wearables data make it possible to address questions that solely EHR-based data sources cannot. Here we provide an overview of the cohort of GLP-1RA users in *All of Us*, demonstrating trends and data availability, to present this platform’s utility for additional GLP-1RA research.

## Methods

This study used data from the Controlled Tier CDRv8 available via the Researcher Workbench. Participants are U.S. residents, aged 18 or older at enrollment, and enrolled between May 6, 2018, and October 1, 2023.

### Defining the GLP-1RA Cohort

Participants with GLP-1RAs were identified via EHR prescription records in the drug exposure table containing OMOP Concept IDs for GLP-1RAs (eTable 1). For this analysis, we used all records in the drug exposure table, including prescriptions written by a clinician, dispensations, claims, administrations, medication list entries, self-reports, and those without a specified source (eTable 2). Here, these records are referred to collectively as “prescriptions.” The GLP-1RA cohort was defined as participants with at least two GLP-1RA prescriptions of any type on different days at any time point.

#### Survey Analysis

We derived demographic and person-level characteristics from surveys participants completed as part of *All of Us*.^6,7^ Surveys include topics such as sociodemographics, physical and behavioral health, and drug use. All participants were required to take the Basics survey, which contains demographic questions. All other surveys are optional or offered to only a subset of the *All of Us* population. We also included items from the Lifestyle and Overall Health surveys. These surveys are optional and offered to all participants but have high response rates in our GLP-1RA cohort (> 98%). We derived sex, race, ethnicity, household income, education, health insurance status and type, employment status, and disability from the Basics survey; cigarette smoking history from the Lifestyle survey; and self-rated health and quality of life from the Overall Health survey.

Counts and proportions for these measures are provided for the GLP-1RA cohort and the *All of Us* CDRv8 population. All proportions are calculated out of the number of individuals who answered each survey, including individuals who selected “prefer not to answer.” As of CDRv8, surveys used in this analysis were offered to participants only once. The *All of Us* Survey Explorer contains additional information on the surveys, questions, and skip patterns.^8^

#### Health Care Utilization Analysis

We classified visits that appeared in the visit occurrence table as inpatient, outpatient, or emergency department (ED) (eTable 3). EHR history was calculated by taking the difference in years between the first and last inpatient, outpatient, or ED visit. We calculated the number of unique inpatient visit days, outpatient visits, and ED visits in each person’s entire EHR, categorized them, and reported counts and proportions for each category.

#### Comorbidity Analysis

We identified select comorbid conditions to examine illness burden among the GLP-1RA cohort. The comorbid condition analysis was limited to conditions identified through EHR data, except body mass index (BMI), which was calculated from self-reported or *All of Us* staff–reported-measurements when available. Participants can have repeated measures; for this study, we report BMI measured closest to the GLP-1RA initiation date for those in our GLP-1RA cohort and to the consent date for the total population.

Authors with clinical expertise selected and reviewed ICD-10-CM and SNOMED CT codes for comorbid conditions (eTable 4). To be counted for each comorbid condition, participants needed at least one record with a code indicating presence of the condition. The code must appear within 24 months before, and any time after, the index date—defined as the first GLP-1RA prescription for the GLP-1RA cohort or the EHR consent date for the CDRv8 population. Reported conditions are those for which GLP-1RAs are an FDA-approved treatment or that current research suggests may be affected by GLP-1RA use. Additionally, we calculated Charlson Comorbidity Index (CCI) scores to characterize chronic disease burden.^9^ We defined the CCI conditions and CCI score using the ICD-9 and -10 codes in Glasheen et al. (eTables 5-7).^10^

#### Prescription Trend Analysis

We examined several GLP-1RA prescription trends among the cohort. The total GLP-1RA treatment period is defined as days from the first recorded GLP-1RA prescription to the last. Treatment episodes are defined as any period with no more than a 90-day gap between prescription records. This allows for 90-day prescriptions and other factors (eg, drug shortages) that might unintentionally interrupt treatment. Treatment gaps are defined as any period longer than 90 days between two GLP-1RA prescriptions.

#### Observations and Measurements Analysis

To explore availability of physical measurements in the EHR data, we looked for BMI, blood pressure, heart rate, and waist and hip circumference values. We also examined availability of laboratory tests for glucose and lipid homeostasis, pancreatic enzymes, liver and kidney function, and other blood tests. Due to the various tests available, we limited the analysis to those with the most common Logical Observation Identifiers Names and Codes (LOINC) code for each condition or any additional code that was present for at least 5% of the CDRv8 population (eTable 8).

#### Wearables Analysis

Fitbit data include daily sleep information, daily step counts, and minute-level heart rate information. For heart rate, we condensed minute-level data into daily average and daily minimum heart rate values. Daily sleep and restless minutes were obtained from the “sleep_daily_summary” table. We used daily step count for the total number of steps recorded per calendar day from the “activity_summary” table. To account for seasonal variability in fitness data,^11,12^ we included only individuals with at least 1 year of GLP-1RA treatment and data present from each month of the year before and after treatment initiation.

### Statistical Methods

Analyses were conducted using a Jupyter Notebook workspace, Python version 3.10.12, and R version 4.3.1 in the Researcher Workbench (https://workbench.researchallofus.org/workspaces/aou-rw-f5da1fd0/glp1raprescriptionusepatternsintheallofusresearchprogram/analysis). We report frequencies and proportions for categorical characteristics and medians with interquartile ranges (IQRs) for continuous characteristics for the GLP-1RA cohort. We provide measures for the entire *All of Us* population for comparison. For the wearables analysis, we performed paired *t*-tests to compare pre- and during-GLP-1RA treatment data for heart rate, sleep, and step count.

## Results

### Data Availability

Of the total CDRv8 population (N=633 534), 15,477 participants (2.4%) have GLP-1RAs on at least two separate days and represent the GLP-1RA cohort described here. Table 1 displays GLP-1RA prescription frequencies in CDRv8. Dulaglutide is the most prevalent GLP-1RA prescription record in *All of Us*, whereas the greatest proportion of people with GLP-1RA prescriptions have at least one prescription for semaglutide. Data availability in *All of Us* is higher across all data types for the GLP-1RA cohort than for the entire CDRv8 population (Table 2). Over 80% of the total CDRv8 population has EHRs available. Because they have been defined through EHRs, the entire GLP-1RA cohort has EHR data, although the availability of distinct data types (eg, procedures, labs) varies. The GLP-1RA cohort has very high data availability across other data types, including near-total representation in physical measurements and several surveys.

**Table 1.** GLP-1RA Prescriptions, by Frequency and Initial Prescribing Date.

| **GLP-1RA type** | **Participants with at least 1 prescription**<br>**N (%)** | **Participants with 2 or more prescription days**<br>**N (%)** | **Total GLP-1RA prescriptions in *All of Us* CDRv8**<br>**N (%)** | **Earliest prescription date** |
| --- | --- | --- | --- | --- |
| Dulaglutide | 8218 (39.6) | 5747 (37.2) | 81 689 (35.4) | 2012-12-26 |
| Semaglutide | 10 461 (50.4) | 7151 (46.3) | 72 126 (31.3) | 2016-06-22 |
| Liraglutide | 6236 (30.0) | 4082 (26.4) | 53 317 (23.1) | 2005-01-01 |
| Exenatide | 1758 (8.5) | 1041 (6.7) | 15 122 (6.6) | 2005-08-05 |
| Albiglutide | 141 (0.7) | 81 (0.5) | 768 (0.3) | 2014-09-04 |
| Lixisenatide | 101 (0.5) | 55 (0.4) | 719 (0.3) | 2017-02-16 |
| Tirzepatide | 1457 (7.0) | 882 (5.7) | 6772 (2.9) | 2022-06-08 |
| Multiple GLP-1RAs | 5968 (28.7) | 3427 (22.2) | NA | 2005-01-01 |
| **Any GLP-1RA** | **20 768 (100.0)** | **15 447 (100.0)<sup>a</sup>** | **230 513 (100.0)** | **2005-01-01** |

Abbreviations: CDRv8, Curated Data Repository version 8; GLP-1RA, glucagon-like peptide-1 receptor agonist.
<sup>a</sup>Analysis cohort.

**Table 2.**
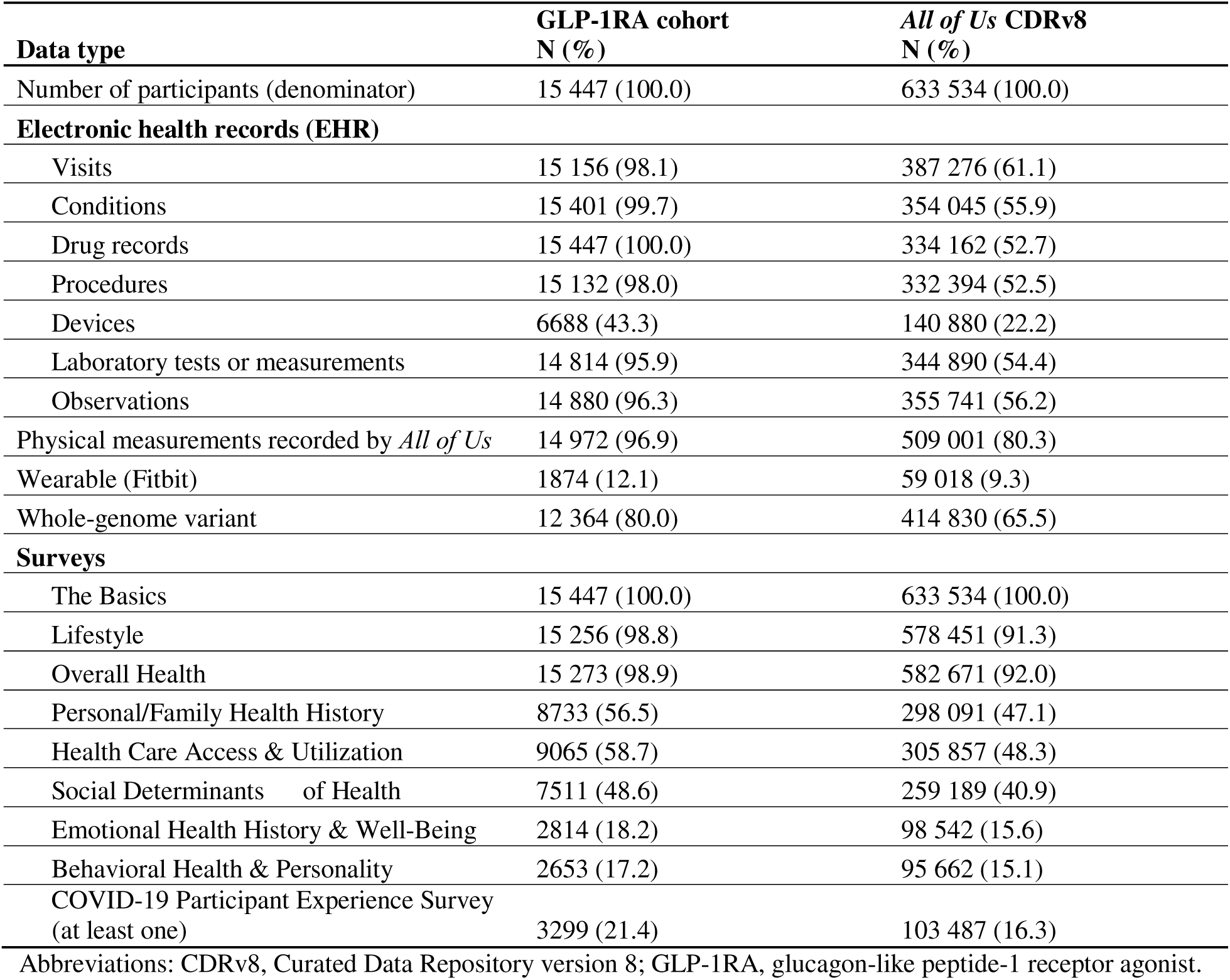
GLP-1RA Cohort *All of Us* CDRv8 Data Availability Summary.

| **Data type** | **GLP-1RA cohort**<br>**N (%)** | ***All of Us* CDRv8**<br>**N (%)** |
| --- | --- | --- |
| Number of participants (denominator) | 15 447 (100.0) | 633 534 (100.0) |
| **Electronic health records (EHR)** |  |  |
| Visits | 15 156 (98.1) | 387 276 (61.1) |
| Conditions | 15 401 (99.7) | 354 045 (55.9) |
| Drug records | 15 447 (100.0) | 334 162 (52.7) |
| Procedures | 15 132 (98.0) | 332 394 (52.5) |
| Devices | 6688 (43.3) | 140 880 (22.2) |
| Laboratory tests or measurements | 14 814 (95.9) | 344 890 (54.4) |
| Observations | 14 880 (96.3) | 355 741 (56.2) |
| Physical measurements recorded by *All of Us* | 14 972 (96.9) | 509 001 (80.3) |
| Wearable (Fitbit) | 1874 (12.1) | 59 018 (9.3) |
| Whole-genome variant | 12 364 (80.0) | 414 830 (65.5) |
| **Surveys** |  |  |
| The Basics | 15 447 (100.0) | 633 534 (100.0) |
| Lifestyle | 15 256 (98.8) | 578 451 (91.3) |
| Overall Health | 15 273 (98.9) | 582 671 (92.0) |
| Personal/Family Health History | 8733 (56.5) | 298 091 (47.1) |
| Health Care Access & Utilization | 9065 (58.7) | 305 857 (48.3) |
| Social Determinants of Health | 7511 (48.6) | 259 189 (40.9) |
| Emotional Health History & Well-Being | 2814 (18.2) | 98 542 (15.6) |
| Behavioral Health & Personality | 2653 (17.2) | 95 662 (15.1) |
| COVID-19 Participant Experience Survey<br>(at least one) | 3299 (21.4) | 103 487 (16.3) |

Abbreviations: CDRv8, Curated Data Repository version 8; GLP-1RA, glucagon-like peptide-1 receptor agonist.

#### Demographics and Survey Responses

GLP-1RA cohort participants are more likely to be female or Black/African American and less likely to be Asian American or Hispanic/Latino than the CDRv8 population (Table 3). The GLP-1RA cohort is generally older, although less likely to be 80 or older, than the CDRv8 population. The GLP-1RA cohort tends to have lower income and education and is more likely to have insurance, particularly Medicare or Medicaid, than the CDRv8 population. Likely owing to their older age, the GLP-1RA cohort is more likely to be disabled or retired, be current or former smokers, and report poorer health and quality of life. Notably, the GLP-1RA cohort appears to have higher health care utilization (inpatient, outpatient, and ED visits) than the CDRv8 population, but they have more EHR data available, which may skew these differences.

**Table 3.**
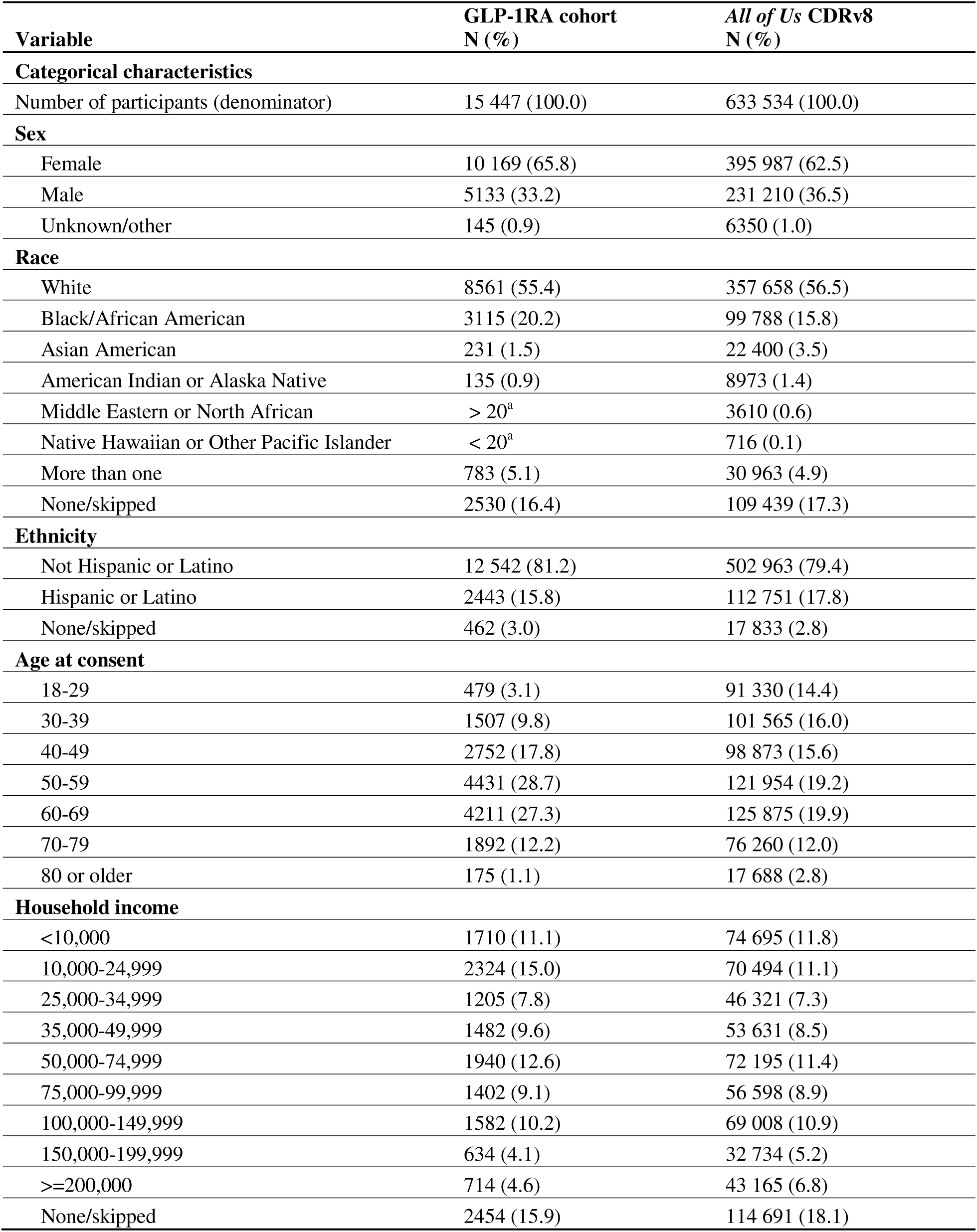
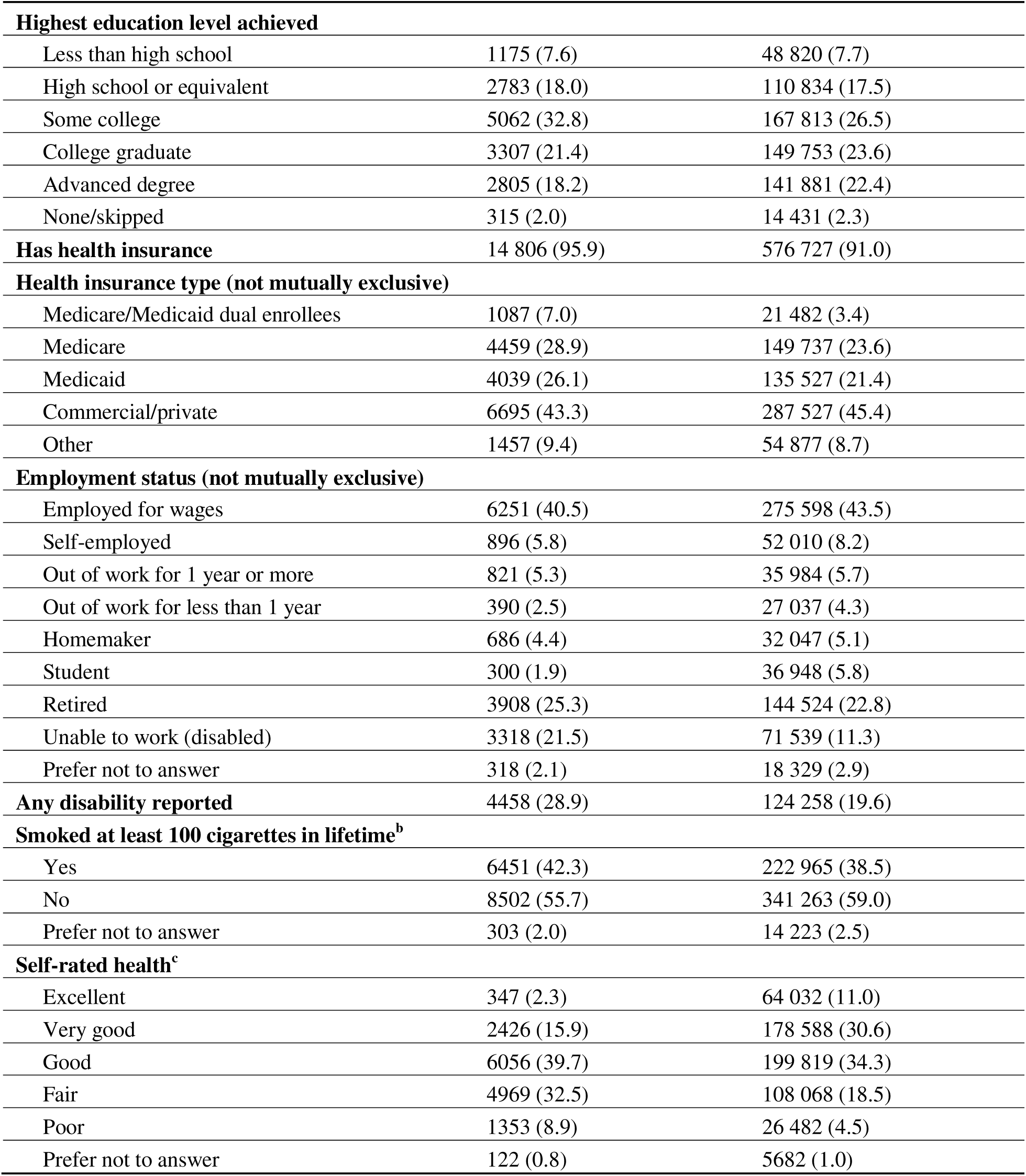
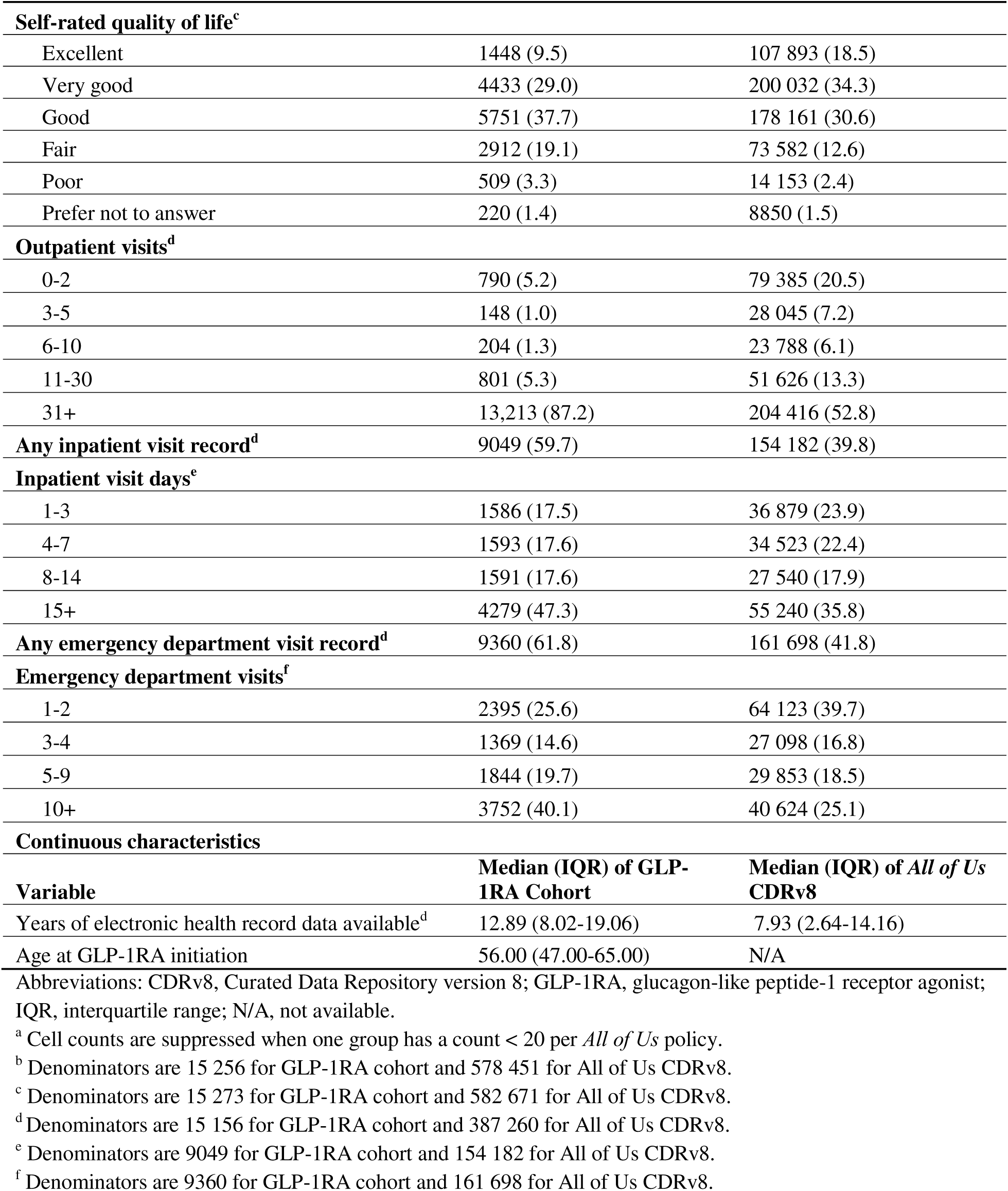
Demographics of GLP-1RA Cohort Compared With *All of Us* CDRv8 Population.

| Variable | GLP-1RA cohort<br>N (%) | *All of Us* CDRv8<br>N (%) |
| --- | --- | --- |
| **Categorical characteristics** |  |  |
| Number of participants (denominator) | 15 447 (100.0) | 633 534 (100.0) |
| **Sex** |  |  |
| Female | 10 169 (65.8) | 395 987 (62.5) |
| Male | 5133 (33.2) | 231 210 (36.5) |
| Unknown/other | 145 (0.9) | 6350 (1.0) |
| **Race** |  |  |
| White | 8561 (55.4) | 357 658 (56.5) |
| Black/African American | 3115 (20.2) | 99 788 (15.8) |
| Asian American | 231 (1.5) | 22 400 (3.5) |
| American Indian or Alaska Native | 135 (0.9) | 8973 (1.4) |
| Middle Eastern or North African | > 20<sup>a</sup> | 3610 (0.6) |
| Native Hawaiian or Other Pacific Islander | < 20<sup>a</sup> | 716 (0.1) |
| More than one | 783 (5.1) | 30 963 (4.9) |
| None/skipped | 2530 (16.4) | 109 439 (17.3) |
| **Ethnicity** |  |  |
| Not Hispanic or Latino | 12 542 (81.2) | 502 963 (79.4) |
| Hispanic or Latino | 2443 (15.8) | 112 751 (17.8) |
| None/skipped | 462 (3.0) | 17 833 (2.8) |
| **Age at consent** |  |  |
| 18-29 | 479 (3.1) | 91 330 (14.4) |
| 30-39 | 1507 (9.8) | 101 565 (16.0) |
| 40-49 | 2752 (17.8) | 98 873 (15.6) |
| 50-59 | 4431 (28.7) | 121 954 (19.2) |
| 60-69 | 4211 (27.3) | 125 875 (19.9) |
| 70-79 | 1892 (12.2) | 76 260 (12.0) |
| 80 or older | 175 (1.1) | 17 688 (2.8) |
| **Household income** |  |  |
| <10,000 | 1710 (11.1) | 74 695 (11.8) |
| 10,000-24,999 | 2324 (15.0) | 70 494 (11.1) |
| 25,000-34,999 | 1205 (7.8) | 46 321 (7.3) |
| 35,000-49,999 | 1482 (9.6) | 53 631 (8.5) |
| 50,000-74,999 | 1940 (12.6) | 72 195 (11.4) |
| 75,000-99,999 | 1402 (9.1) | 56 598 (8.9) |
| 100,000-149,999 | 1582 (10.2) | 69 008 (10.9) |
| 150,000-199,999 | 634 (4.1) | 32 734 (5.2) |
| >=200,000 | 714 (4.6) | 43 165 (6.8) |
| None/skipped | 2454 (15.9) | 114 691 (18.1) |

**Table 3.** Demographics of GLP-1RA Cohort Compared With *All of Us* CDRv8 Population (continued)
| **Variable** | **GLP-1RA cohort**<br>**N (%)** | ***All of Us* CDRv8**<br>**N (%)** |
| --- | --- | --- |
| **Highest education level achieved** |  |  |
| Less than high school | 1175 (7.6) | 48 820 (7.7) |
| High school or equivalent | 2783 (18.0) | 110 834 (17.5) |
| Some college | 5062 (32.8) | 167 813 (26.5) |
| College graduate | 3307 (21.4) | 149 753 (23.6) |
| Advanced degree | 2805 (18.2) | 141 881 (22.4) |
| None/skipped | 315 (2.0) | 14 431 (2.3) |
| **Has health insurance** | 14 806 (95.9) | 576 727 (91.0) |
| **Health insurance type (not mutually exclusive)** |  |  |
| Medicare/Medicaid dual enrollees | 1087 (7.0) | 21 482 (3.4) |
| Medicare | 4459 (28.9) | 149 737 (23.6) |
| Medicaid | 4039 (26.1) | 135 527 (21.4) |
| Commercial/private | 6695 (43.3) | 287 527 (45.4) |
| Other | 1457 (9.4) | 54 877 (8.7) |
| **Employment status (not mutually exclusive)** |  |  |
| Employed for wages | 6251 (40.5) | 275 598 (43.5) |
| Self-employed | 896 (5.8) | 52 010 (8.2) |
| Out of work for 1 year or more | 821 (5.3) | 35 984 (5.7) |
| Out of work for less than 1 year | 390 (2.5) | 27 037 (4.3) |
| Homemaker | 686 (4.4) | 32 047 (5.1) |
| Student | 300 (1.9) | 36 948 (5.8) |
| Retired | 3908 (25.3) | 144 524 (22.8) |
| Unable to work (disabled) | 3318 (21.5) | 71 539 (11.3) |
| Prefer not to answer | 318 (2.1) | 18 329 (2.9) |
| **Any disability reported** | 4458 (28.9) | 124 258 (19.6) |
| **Smoked at least 100 cigarettes in lifetime<sup>b</sup>** |  |  |
| Yes | 6451 (42.3) | 222 965 (38.5) |
| No | 8502 (55.7) | 341 263 (59.0) |
| Prefer not to answer | 303 (2.0) | 14 223 (2.5) |
| **Self-rated health<sup>c</sup>** |  |  |
| Excellent | 347 (2.3) | 64 032 (11.0) |
| Very good | 2426 (15.9) | 178 588 (30.6) |
| Good | 6056 (39.7) | 199 819 (34.3) |
| Fair | 4969 (32.5) | 108 068 (18.5) |
| Poor | 1353 (8.9) | 26 482 (4.5) |
| Prefer not to answer | 122 (0.8) | 5682 (1.0) |

**Table 3.** Demographics of GLP-1RA Cohort Compared With *All of Us* CDRv8 Population (continued)
| Variable | GLP-1RA cohort<br>N (%) | *All of Us* CDRv8<br>N (%) |
| --- | --- | --- |
| **Self-rated quality of life<sup>c</sup>** |  |  |
| Excellent | 1448 (9.5) | 107 893 (18.5) |
| Very good | 4433 (29.0) | 200 032 (34.3) |
| Good | 5751 (37.7) | 178 161 (30.6) |
| Fair | 2912 (19.1) | 73 582 (12.6) |
| Poor | 509 (3.3) | 14 153 (2.4) |
| Prefer not to answer | 220 (1.4) | 8850 (1.5) |
| **Outpatient visits<sup>d</sup>** |  |  |
| 0-2 | 790 (5.2) | 79 385 (20.5) |
| 3-5 | 148 (1.0) | 28 045 (7.2) |
| 6-10 | 204 (1.3) | 23 788 (6.1) |
| 11-30 | 801 (5.3) | 51 626 (13.3) |
| 31+ | 13,213 (87.2) | 204 416 (52.8) |
| **Any inpatient visit record<sup>d</sup>** | 9049 (59.7) | 154 182 (39.8) |
| **Inpatient visit days<sup>e</sup>** |  |  |
| 1-3 | 1586 (17.5) | 36 879 (23.9) |
| 4-7 | 1593 (17.6) | 34 523 (22.4) |
| 8-14 | 1591 (17.6) | 27 540 (17.9) |
| 15+ | 4279 (47.3) | 55 240 (35.8) |
| **Any emergency department visit record<sup>d</sup>** | 9360 (61.8) | 161 698 (41.8) |
| **Emergency department visits<sup>f</sup>** |  |  |
| 1-2 | 2395 (25.6) | 64 123 (39.7) |
| 3-4 | 1369 (14.6) | 27 098 (16.8) |
| 5-9 | 1844 (19.7) | 29 853 (18.5) |
| 10+ | 3752 (40.1) | 40 624 (25.1) |
| **Continuous characteristics** |  |  |
| Variable | Median (IQR) of GLP-1RA Cohort | Median (IQR) of *All of Us* CDRv8 |
| Years of electronic health record data available<sup>d</sup> | 12.89 (8.02-19.06) | 7.93 (2.64-14.16) |
| Age at GLP-1RA initiation | 56.00 (47.00-65.00) | N/A |

Abbreviations: CDRv8, Curated Data Repository version 8; GLP-1RA, glucagon-like peptide-1 receptor agonist; IQR, interquartile range; N/A, not available.
<sup>a</sup> Cell counts are suppressed when one group has a count < 20 per *All of Us* policy.
<sup>b</sup> Denominators are 15 256 for GLP-1RA cohort and 578 451 for *All of Us* CDRv8.
<sup>c</sup> Denominators are 15 273 for GLP-1RA cohort and 582 671 for *All of Us* CDRv8.
<sup>d</sup> Denominators are 15 156 for GLP-1RA cohort and 387 260 for *All of Us* CDRv8.
<sup>e</sup> Denominators are 9049 for GLP-1RA cohort and 154 182 for *All of Us* CDRv8.
<sup>f</sup> Denominators are 9360 for GLP-1RA cohort and 161 698 for *All of Us* CDRv8.

#### Comorbid Conditions

Unsurprisingly, the GLP-1RA cohort has a much higher prevalence of type 2 diabetes, obesity, and severe obesity than the CDRv8 population and is over 5 times more likely to have concurrent type 2 diabetes and obesity (Table 4). Notably, about 20% of *All of Us* participants with type 2 diabetes and obesity are in the GLP-1RA cohort. The GLP-1RA cohort also has a higher prevalence of nearly every other condition assessed except alcohol use disorder. The GLP-1RA cohort also had much higher CCI scores. Over 10% of CDRv8 participants with a CCI score greater than 5 were in the GLP-1RA cohort.

**Table 4.** Comorbidities Present in the Electronic Health Record for the GLP-1RA Cohort Compared With *All of Us* CDRv8 Population.

| Comorbidities | GLP-1RA cohort<br>N (%) | *All of Us* CDRv8<br>N (%) |
| --- | --- | --- |
| Type 2 diabetes | 11 511 (74.5) | 68 576 (17.4) |
| Weight |  |  |
| Overweight (BMI: 25–29.9) | 2070 (13.4) | 155 909 (30.4) |
| Obese (BMI: 30–39.9) | 7835 (50.8) | 162 619 (31.7) |
| Severe obesity (BMI: 40+) | 5108 (33.1) | 49 506 (9.7) |
| Both type 2 diabetes and obesity (BMI: $\geq 30$) | 9473 (61.3) | 43 600 (11.1) |
| Obstructive sleep apnea | 6574 (42.6) | 51 816 (13.2) |
| Type 1 diabetes | 1179 (7.6) | 5792 (1.5) |
| Hypertension | 13 770 (89.1) | 226 329 (57.5) |
| Hyperlipidemia | 11 409 (73.9) | 137 107 (34.8) |
| Chronic kidney disease | 3572 (23.1) | 35 406 (9.0) |
| Metabolic dysfunction-associated steatotic liver disease | 3170 (20.5) | 23 361 (5.9) |
| Alcohol-associated liver disease | 137 (0.9) | 3155 (0.8) |
| Alcohol use disorder | 719 (4.7) | 19 353 (4.9) |
| Substance use disorder | 3313 (21.4) | 68 602 (17.4) |
| Depression | 7553 (48.9) | 98 911 (25.1) |
| Anxiety | 6801 (44.0) | 99 652 (25.3) |
| Alzheimer’s disease | 63 (0.4) | 1086 (0.3) |
| Parkinson’s disease | 141 (0.9) | 2263 (0.6) |
| Sarcopenia | < 20<sup>a</sup> | 148 (0.0) |
| Charlson Comorbidity Index |  |  |
| 0 | 2497 (16.2) | 214 588 (54.5) |
| 1-2 | 5075 (32.9) | 103 512 (26.3) |
| 3-4 | 3961 (25.6) | 40 422 (10.3) |
| 5+ | 3914 (25.3) | 35 074 (8.9) |

Abbreviations: BMI, body mass index; CDRv8, Curated Data Repository version 8; GLP-1RA, glucagon-like peptide-1 receptor agonist.
<sup>a</sup> Counts fewer than 20 are suppressed per *All of Us* policy.

#### Prescription Trends

GLP-1RA prescriptions are present for our cohort every year from 2005 to 2023. The median length between first and last GLP-1RA prescription is 1.5 years (IQR 0.6–3.6). Participants have a median of two distinct episodes (ie, series of prescriptions with no more than 90 days between start dates) of GLP-1RA prescriptions (IQR 1–4). Treatment episodes last a median of 84 days (IQR 41–175). Within treatment episodes of two or more prescriptions, the median time between prescriptions is 31 days (IQR 21.5–52). Among this cohort, 10 697 participants (69.2%) have a treatment gap of 90 days or greater. For these participants, there is a median of one treatment gap (IQR 1–2). The median gap length (ie, time between episodes) is 169 days (IQR 115–308).

GLP-1RA prescriptions increase year over year for the entire period of data availability. The cohort includes fewer than 20 participants and fewer than 20 unique prescriptions in 2005, when FDA first approved exenatide to treat diabetes, and increases to 605 participants with 2413 prescriptions in 2014, when FDA first approved liraglutide to treat weight loss. In 2022, the last full year of data, the cohort includes 9331 participants who had 43,266 GLP-1RA prescriptions. Figure 1 shows counts of prescriptions by each GLP-1RA. Most GLP-1RA prescriptions from 2005 to 2012 are for exenatide. In 2013, liraglutide becomes the most common GLP-1RA. Dulaglutide becomes increasingly common starting in 2015, semaglutide increases beginning in 2018, and tirzepatide appears in 2022. By 2022, 45.5% of prescriptions are semaglutide, followed by dulaglutide (39.4%), liraglutide (11.2%), tirzepatide (3.1%), exenatide (0.6%), and lixisenatide (0.2%).

**Figure 1.**
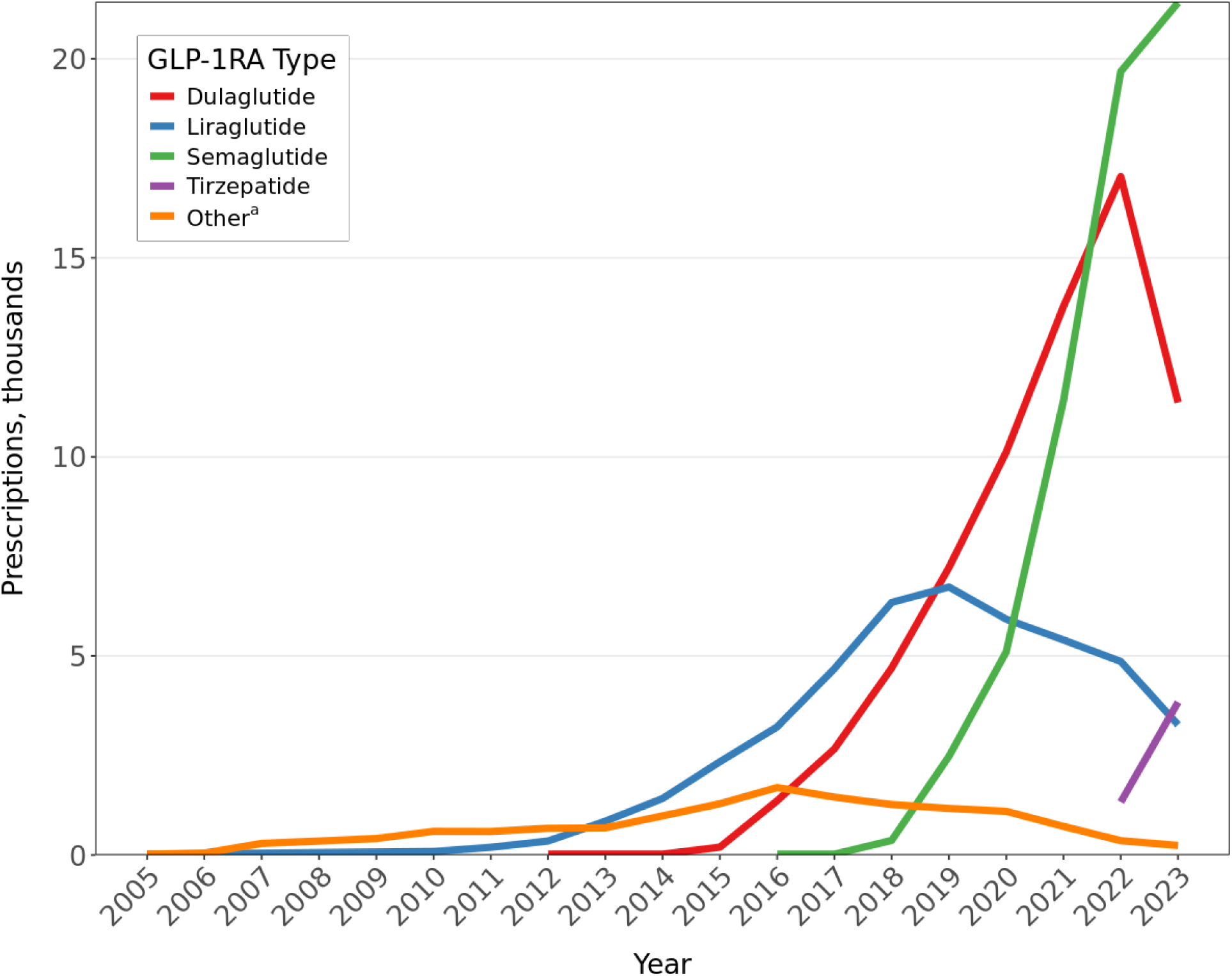
Annual Number of GLP-1RA Prescriptions Among *All of Us* Participants Abbreviation: GLP-1RA, glucagon-like peptide-1 receptor agonist. ^a^Includes albiglutide, exenatide, and lixisenatide.

#### Observations and Measurements

Physical measurements and laboratory test results are available for most of the GLP-1RA cohort. Most of these participants have at least one measurement available for BMI (99.6%), systolic and diastolic blood pressure (99.4%), heart rate (93.1%), waist circumference (81.7%), and hip circumference (81.6%). Among common laboratory tests, there is extensive coverage for participants with at least one glycated hemoglobin (89.5%) and lipid profile, including high-density lipoprotein (88.2%), low-density lipoprotein (87.5%), total cholesterol (88.5%), and triglycerides (85.5%). A total of 40.9% of the participants have C-reactive protein results. Most participants have at least one liver test, including alanine transaminase (91.9%), alkaline phosphatase (90.9%), aspartate aminotransferase (90.6%), bilirubin in blood (88.8%), bilirubin in urine (64.1%), and gamma-glutamyl transferase (15.7%). Blood counts are common, including hemoglobin (92.4%), red blood cell (86.3%), platelet (85.3%), and white blood cell (84.6%). Creatinine in blood (94.5%) and urine (64.6%) and calculated estimated glomerular filtration rate (76.0%) also are widely available. Pancreas tests are less common, including results for lipase (43.6%) and amylase (22.1%).

#### Fitbit Data

Fitbit data are present for 1874 individuals (12%) in our GLP-1RA cohort. Heart rate parameters increased after initiating GLP-1RA prescriptions, as has been reported in other literature.^13,14^ Daily minimum heart rate increased by 1.77 beats per minute (95% confidence interval: 0.64, 2.90; *P* = 0.0027). Daily average heart rate, daily step count, daily sleep minutes, and daily restless minutes processed in the same manner did not show statistically significant changes between pre- and post-GLP-1RA initiation periods (eTable 9).

## Discussion

Using *All of Us* CDRv8 EHR data, we identified a cohort of 15,447 participants with two or more GLP-1RA prescriptions on separate days to assess demographics, comorbidities, prescription trends, activity patterns, and data availability. *All of Us* GLP-1RA prescription trends are like those reported in other large EHR data systems.^15^ This cohort is diverse along several sociodemographic features but tends to be older, sicker, more likely to have insurance coverage, and more likely to use health care than the average *All of Us* participant. Various data types are available for participants with GLP-1RA prescriptions, including numerous survey responses and long EHR histories that include a wealth of information on prescriptions, comorbid conditions, physical measurements, and laboratory tests. For example, our brief exploration of the Fitbit data available in *All of Us* shows increases in heart rate after starting GLP-1RA treatment, in line with earlier research.^16^ This exploration of people prescribed GLP-1RAs demonstrates that *All of Us* can be a powerful data source to answer questions about real-world GLP-1RA use across various demographics and outcomes.

Given the longitudinal and multimodal nature of its data, *All of Us* provides insights into GLP-1RA users that other data sources cannot. In addition to clinical findings, *All of Us* can characterize associations between GLP-1RAs and mental health, behavior, physical activity, and genetics. Survey data provide a breadth of demographic, attitudinal, and behavioral information unavailable in most observational EHR-based research. Fitbit data, likewise, provide information on physical activity that is rarely available in EHR data outside specific research contexts. Because *All of Us* purposely oversamples groups with limited health outcome data, it presents opportunities to study GLP-1RA use among populations that are often less prevalent in other studies, including communities with barriers that prevent them from accessing treatments such as GLP-1RAs.

### Limitations

Using *All of Us* data to study GLP-1RA use has several limitations. Notably, *All of Us* targeted recruitment of participants unsystematically and therefore is not representative of the U.S. adult population.^17^ *All of Us* participants may be more willing than the average person to be involved in a large national study, respond to surveys, submit biomedical samples, or consent to providing health data. As a result, we do not recommend making direct comparisons of these findings with national and international estimates.

Other limitations are common to observational research using EHR data.^18,19^ Participants with EHR data are, by definition, able to access health care resources, potentially introducing selection bias. EHR data may be incomplete, particularly if a participant has a fragmented digital medical history across various health care providers, and *All of Us* may not have access to data from all of a participant’s providers. Additionally, EHRs can more easily identify a condition’s presence than its resolution or absence. The data available in *All of Us* do not include rule-out diagnoses, and CDRv8 does not currently include clinical notes.

With prescription data in *All of Us* from the EHR data alone, it can be difficult to identify when a prescription was dispensed to a participant and impossible to know whether the participant administered it. Many GLP-1RAs are still relatively new to the market, and trends in use may significantly change. Additionally, GLP-1RA shortages during the data availability period could have caused participants to seek the drugs through means EHRs cannot reliably capture.^20^

### Future Directions

Multimodal data availability in *All of Us* allows for a wide array of research opportunities. Longitudinal EHRs combined with survey, behavioral, genomic, and wearables data are typically not available at this scale in most studies. These data can support studies of emerging and off-label uses of GLP-1RAs. For example, alcohol and other substance use disorders, mental health, and neurodegenerative diseases have been early areas of research.^21,22^ The data can also be used to better understand emerging trends such as switching between GLP-1RA types as insurance coverage has shifted. The anticipated release of CDRv9 will include additional data through December 2024, with an expected increase in the number of GLP-1RA prescriptions.

## Data Availability

The Featured Workspace is hosted on the All of Us Researcher Workbench and contains code and data used for the analysis in the paper https://workbench.researchallofus.org/workspaces/aou-rw-f5da1fd0/glp1raprescriptionusepatternsintheallofusresearchprogram/analysis

## Author Contributions

Ms. Gasdaska, Drs. Tyndall and Preble, and Mr. Brannock had full access to all data in the study and take responsibility for the integrity of the data and the accuracy of the data analysis.

*Concept and design:* All authors

*Acquisition, analysis, or interpretation of data:* All authors

*Drafting of the manuscript:* Gasdaska, Tyndall, Preble, Brannock, Huda

*Critical review of the manuscript for important intellectual content:* All authors

*Statistical analysis:* Gasdaska, Tyndall, Preble, Brannock

*Obtained funding:* Adjemian

*Administrative, technical, or material support:* Litwin, Sastry, Adjemian

*Supervision:* McPheeters, Marcial, Adjemian

## Additional Contributions

We gratefully acknowledge *All of Us* participants for their contributions, without whom this research would not have been possible. We also thank the National Institutes of Health’s *All of Us* Research Program for making available the participant data examined in this study.

We thank Sheryl Cates for project administration and Jennifer D. Uhrig, PhD, and Megan A. Lewis, PhD, the award multiple principal investigators, for supporting this project (RTI International).

## Data and code Availability

The Featured Workspace is hosted on the *All of Us* Researcher Workbench and contains code and data used for the analysis in the paper https://workbench.researchallofus.org/workspaces/aou-rw-f5da1fd0/glp1raprescriptionusepatternsintheallofusresearchprogram/analysis

