## Supplemental Materials for "GLP-1 Receptor Agonist Prescription Patterns in the *All of Us* Research Program"

**Supplemental Material**

**eTable 1.** OMOP Concept IDs and RxNorm codes for GLP-1RA drugs

**eTable 2.** Drug exposure types for all CDRv8 GLP-1RA exposures

**eTable 3.** Inpatient, outpatient, or emergency department (ED) concepts

**eTable 4.** Condition-related OMOP Concept IDs

**eTable 5.** Charlson Comorbidity Index scoring system

**eTable 6.** Charlson Comorbidity Index hierarchy categories

**eTable 7.** ICD-9 and ICD-10 codes for Charlson Comorbidity Index conditions

**eTable 8.** Laboratory tests and LOINC or SNOMED CT codes

**eTable 9.** Fitbit wearable sensor data analysis

Curated Data Repository version 8 (CDRv8) electronic health record (EHR) data conform to the Observational and Medical Outcomes Partnership (OMOP) common data model, which provides a standardized data structure for EHR data from multiple health care providers and systems. Statistical code and analytic approaches adopted for other OMOP projects can therefore be used with *All of Us* data. OMOP’s vocabulary and ontology hierarchy provide a way to quickly aggregate records that use semantically similar concepts coded under different terminologies or ontologies (eg, by relating International Classification of Diseases version 10, Clinical Modification [ICD-10-CM] and Systematized Nomenclature of Medicine Clinical Terms [SNOMED CT] codes for the same condition). OMOP Concept IDs used in this paper are provided in eTables 1, 3, and 4.

We collapsed some response categories for selected survey questions. We collapsed the responses to the educational attainment question (never attended, grades 1-4, grades 5-8, and grades 9-11) into a “less than high school” category. For health insurance, we collapsed “insurance purchased directly from an insurance company” and “insurance through a current or former employer or union” into a “commercial/private” category. We also combined TRICARE, Veterans Affairs, Indian Health Service, and “any other type of health insurance” responses into a single “other” category. Finally, we reported participants as having a disability if they answered yes to any one of the six disability questions in the Basics survey.^1^

EHR code sets were derived from ICD-10-CM and SNOMED CT codes in selected value sets in the National Library of Medicine’s Value Set Authority Center, the Center for Medicare & Medicaid Services Chronic Conditions Warehouse, and codes used to construct the Charlson and Elixhauser Comorbidity Indices, and through reviews of literature for certain conditions.

**eTable 1. OMOP Concept IDs and RxNorm Codes for GLP-1RA Drugs**

| **GLP-1RA** | **OMOP Concept ID** | **RxNorm code** |
| --- | --- | --- |
| Albiglutide^a^ | 44816332 | 1534763 |
| Dulaglutide | 45774435 | 1551291 |
| Exenatide | 1583722 | 60548 |
| Liraglutide | 40170911 | 475968 |
| Lixisenatide | 44506754 | 1440051 |
| Semaglutide | 793143 | 1991302 |
| Tirzepatide^b^ | 779705 | 2601723 |

Abbreviations: GLP-1RA, glucagon-like peptide-1 receptor agonist; OMOP, Observational and Medical Outcomes Partnership.

^a^Withdrawn worldwide in 2018.

^b^Dual GLP-1RA and glucose-dependent insulinotropic polypeptide (GIP) receptor agonist.

**eTable 2. Drug Exposure Types^a^ for All CDRv8 GLP-1RA Exposures**

| **Record type** | **Count** |
| --- | --- |
| Prescription dispensed/pharmacy claim | 91 796 |
| Prescription written | 82 075 |
| Medication list entry | 37 360 |
| Other/not specified | 17 099 |
| Drug administration | 1724 |
| Self-report | 459 |
| **Grand total** | **230** **513** |

Abbreviations: CDRv8, Curated Data Repository version 8; GLP-1RA, glucagon-like peptide-1 receptor agonist.

^a^Drug exposure types were reclassified to combine similar categories (e.g., “Medication list entry” and “EHR medication list” were reclassified as “Medication list entry”).

**eTable 3. Inpatient, Outpatient, and Emergency Department Concept IDs**

| **Visit type** | **OMOP Concept IDs** |
| --- | --- |
| Inpatient visits | 42898160, 8920, 38004311, 38004284, 38004303, 8971, 8546, 32037, 8676, 38004515, 38004285, 262, 581379, 8717, 4313303, 9201 |
| Outpatient visits | 38004250, 38004251, 38004207, 8756, 38004258, 581475, 38004249, 38004696, 8964, 8966, 38004245, 581476, 38004218, 38004228, 38004238, 581479, 38004259, 43527986, 38004264, 8949, 38004222, 43527904, 38004269, 38004208, 38004225, 8883, 581385, 38004247, 38004262, 5083, 38004268, 38004267, 38004519, 38004677, 8782, 581477, 8947, 38004246, 38004227, 9202 |
| Emergency department visits | 4163685, 9203, 8870 |

Abbreviations: OMOP, Observational and Medical Outcomes Partnership.

**eTable 4. Condition-Related OMOP Concept IDs**

| Condition | OMOP Concept IDs |
| --- | --- |
| Alzheimer’s disease | 378419, 4220313, 4218017, 4278830, 43530664, 4277444, 44782432, 44784643, 37117145, 4043379 |
| Anxiety | 442077, 434613, 436676, 4211231, 440083, 36684319, 436075, 443414, 4178114, 440374, 4338031, 4058397, 440690, 4304010, 381537, 4147466, 4009184, 436074, 4195865, 443876, 4321835, 4100536, 4146660, 434920, 4084868, 4219627, 436959, 4053320, 4039212, 4268775, 4306640, 4221077, 4189538, 4182847, 4056690, 433178, 4199892, 4198826, 434628, 4242095, 4178928, 35615153, 439786, 4138454, 40484109, 37117155, 35615152, 35615155, 4035299, 763092, 437537, 4252239, 4103276, 4204708, 42538592, 4277739, 4220989, 37110565, 4218624, 4246992, 4217601 |
| Alcohol-associated liver disease | 196463, 46269816, 4340383, 193256, 201612, 201343, 46269835, 4340386, 4340385, 46269818 |
| Alcohol use disorder | 433753, 435243, 4104431, 375519, 196463, 436953, 4205002, 4340383, 193256, 201612, 4340493, 377830, 4310679, 4340964, 376383, 437257, 201343, 435532, 4088373, 4340386, 4146660, 4050977, 439277, 4218106, 4109691, 372607, 375794, 45757093, 374317, 37017563, 441261, 378726, 37016267, 37018356, 435534, 374623, 36713086, 4340385, 440685, 442582, 4078688, 4152165, 40483827, 37016173, 4302744, 4288013, 37016266, 4028805, 4204015 |
| Chronic kidney disease | 46271022, 443597, 44782429, 43531578, 45768812, 443601, 45763854, 439696, 443612, 193782, 44784439, 45763855, 198124, 443919, 44784621, 443611, 443614, 44782690, 44784638, 44782728, 4030520, 43021852, 43020455, 198185, 36716947, 45771064, 45773688, 439695, 439694, 443961, 4128067, 37018886, 43531653, 37019193, 37398911, 4128221, 45769905, 37017813, 4030664, 43531577, 37017104, 44782689, 45768813, 44782691, 4125970, 37395652, 45771075, 43021835, 45757499, 44784639, 45757535, 45769906, 43531559, 46273164, 760850, 43021748, 43531566, 44782692, 45773576, 43531562, 37018761, 4206115, 45757446, 45757445, 45769901, 45769903, 43020457, 45771067, 46273636, 4221487 |

(continued)

**eTable 4. Condition-Related OMOP Concept IDs (continued)**

| Condition | OMOP Concept IDs |
| --- | --- |
| Depression | 440383, 4282096, 4077577, 433440, 436665, 4282316, 444100, 442306, 36684319, 44784632, 435220, 432883, 4228802, 4049623, 4195572, 439254, 438998, 441534, 432876, 4307956, 4338031, 434911, 4098302, 4244078, 4205002, 437528, 444038, 440078, 438406, 4239471, 439253, 432866, 4224940, 4152280, 443906, 435226, 439248, 443237, 4242733, 4328217, 4150985, 439256, 433992, 432285, 35622934, 439249, 442570, 441834, 4012869, 4314692, 4149320, 439246, 4105930, 372599, 440696, 443797, 436386, 439251, 4151170, 438727, 4092239, 40481798, 43531624, 439255, 37018688, 37396201, 4298317, 4329560, 4149321, 441836, 36717092, 4103574, 36713698, 4154309, 437249, 607543, 4307111, 435520, 35610112, 35624743, 4103853, 42872411, 4280361, 4037669, 4102936, 4333678, 440980, 4307804, 4220617, 37016718, 35624748, 42872722, 437250, 439259, 4287544, 4336957, 440067, 432290, 4030856, 35624747, 42872413, 433751, 4215917, 438119, 4141292, 439785, 4094358, 4250023, 439250, 443864, 4172156, 35624744, 439272, 35615153, 4029464, 440698, 4028027, 4269143, 438405, 439273, 607540, 4324945, 37111697, 440079, 35624745, 4327337, 4262111, 44782943, 35615152, 4114950, 37109941, 439262, 4102603, 439001, 4155798, 4241158, 42538589, 373176, 36714998, 4175329, 4205471, 37018689, 4185096, 4057218, 4174987, 4223090, 37312578, 4144519, 43020451, 37016268, 36714389, 4001733, 4071442, 4161200, 4154283, 4304140, 762060, 36715000, 4237734, 4129184, 4031328, 4144233, 37110429, 42538590, 4307951, 4243308, 4096229, 4133073, 4226155, 4307518, 4141603, 4197669, 4154391, 437532, 4154805, 4103126, 4148934, 4217940, 4224639 |
| Hyperlipidemia | 432867, 437827, 438720, 440360, 4029305, 4134862, 4120314, 4047805, 437521, 40483758, 4030619, 45770880, 4142496, 43531651, 4292672, 4079876, 4029260, 4104485, 4031945, 45757432, 40483651, 4299409, 4077382, 4030618, 4029261, 4079886, 43530660, 4144529, 4029892, 4029259, 4220010 |
| Hypertension | 320128, 312648, 44782429, 444101, 316866, 439696, 319034, 44784439, 443919, 319826, 44784621, 376965, 317898, 4110948, 44782690, 313502, 314378, 4028741, 44784638, 312938, 442604, 44782728, 439698, 314369, 195556, 43021852, 43020455, 442626, 314958, 45771064, 439695, 442603, 439694, 4289933, 316994, 37018886, 4263504, 318437, 37208172, 4209293, 4322893, 4167358, 44782689, 37311148, 44782691, 4159755, 4049389, 43021835, 45757140, 4276511, 46273164, 4058987, 44784639, 760850, 44782692, 4183981, 4263067, 43020457, 4135463, 4178312, 4322735, 45757445, 45757446, 4180283, 45771067, 42538697, 4262182, 46273636 |
| Metabolic dysfunction-associated steatotic liver disease | 4059290, 201613, 40484532, 4026131, 40482277, 36716710 |
| Obstructive sleep apnea | 442588, 42872389, 4043564 |
| Parkinson’s disease | 381270, 4196433, 37395785 |
| Sarcopenia | 36674346 |

(continued)

**eTable 4. Condition-Related OMOP Concept IDs (continued)**

| Condition | OMOP Concept IDs |
| --- | --- |
| Substance use disorder | 4209423, 437264, 433753, 435243, 4300092, 438120, 4239381, 434327, 432303, 438130, 4032799, 4104431, 40479573, 4004672, 4264766, 4178114, 375519, 4080762, 436389, 4336384, 440387, 36714559, 196463, 440693, 40483172, 440069, 441260, 433735, 436953, 4205002, 436370, 444038, 443236, 4193868, 4340383, 607217, 436079, 4146763, 193256, 201612, 439796, 4340493, 442601, 377830, 4310679, 433994, 4192127, 4340964, 376383, 375504, 4101256, 436089, 4067987, 4052690, 437257, 201343, 4012869, 435532, 4233811, 435792, 43020446, 4088373, 433452, 440987, 4340386, 4105930, 4102817, 4099809, 373449, 44782714, 437245, 440692, 4146660, 40482898, 433746, 4050977, 439277, 4176464, 435533, 4216493, 4203152, 442915, 443559, 4218106, 4109691, 440379, 372607, 375794, 43021844, 45757093, 4221077, 4191592, 4197130, 374317, 433180, 4002572, 4290538, 437838, 607209, 37017563, 37110444, 4272033, 440992, 4199769, 4100520, 44783367, 441261, 378726, 4103413, 440380, 4173746, 4155336, 37018356, 4056690, 435534, 374623, 4141524, 434016, 607208, 37309681, 45766641, 36713086, 4103853, 4290062, 4148140, 434627, 4137236, 443930, 4198826, 4340385, 44784625, 4097389, 434019, 4184438, 4024296, 440685, 442582, 4272313, 4152165, 4078688, 4230779, 4264889, 37119151, 437533, 4338026, 434328, 4099811, 438393, 4220197, 4099935, 4333676, 443534, 4332883, 37110468, 432878, 37311993, 4176120, 4236877, 4029464, 4239812, 44784627, 37205764, 45773120, 4331287, 42539146, 442913, 37110437, 42872387, 40483827, 4323580, 37108774, 439313, 4307098, 4313135, 4137955, 4035299, 4171175, 42537772, 439554, 42538589, 4176286, 607660, 4009647, 37109022, 42538586, 435798, 4262566, 4302744, 37018689, 432304, 42539702, 440381, 442914, 4232492, 42539355, 37016268, 42538592, 436371, 438732, 4136662, 4332882, 4197434, 4028641, 4204015, 4199244, 42538590, 37110436, 37110407, 4300655, 4103419, 3654785, 4102821, 4028805, 4139421, 37108712, 37110474, 4319464, 4327348, 4333674, 37209507, 4163514, 42537692, 438126, 433745, 37016266, 37110408, 4091715, 43020430, 45766642, 4332990 |
| Type 1 diabetes | 443412, 201254, 37016348, 40484648, 200687, 377821, 45769876, 435216, 37017431, 42538169, 4227210, 37016767, 318712, 45763584, 37016179, 439770, 45770902, 43531008, 45769832, 4225656, 37016180, 4099214, 45769830, 4223303, 4145827, 4225055, 45773688, 45763583, 4224254, 4228112, 4152858, 4063042, 37312218, 201531, 45769873, 609120, 45757674, 35626069, 443592, 609098, 37018566, 609118, 4143857, 609115, 45757507, 45757362, 609100, 45757432, 36713094, 603320, 4047906, 43531009, 603319, 609102, 37017429, 603318, 45771075, 37016353, 37312201, 45773576, 43530660, 45757535, 43531006, 609070, 45771067, 37312200, 4102018, 609111, 43531565, 609110, 45757074, 45769901, 45769903, 4099215 |

(continued)

**eTable 4. Condition-Related OMOP Concept IDs (continued)**

| Condition | OMOP Concept IDs |
| --- | --- |
| Type 2 diabetes | 4193704, 201826, 37016349, 376065, 43531578, 37017432, 443729, 443731, 443732, 45757363, 443733, 45757435, 43530690, 4221495, 37016768, 40482801, 43531616, 43530685, 4099651, 45770830, 45770881, 4222876, 43531563, 4222415, 443734, 4130162, 43530656, 37016354, 45757474, 4196141, 201530, 4226121, 4304377, 4200875, 43531564, 36714116, 4140466, 45773064, 45770880, 4063043, 45771064, 35626070, 609103, 43530689, 43531653, 4228443, 45757449, 4230254, 43531651, 36712687, 609117, 609099, 609106, 36712686, 609101, 4099216, 45769905, 37312204, 609119, 609116, 4215719, 43531577, 37018728, 4177050, 609104, 4129519, 609109, 609105, 602345, 37312205, 43531597, 46274058, 45757278, 45769888, 45757499, 609112, 45757277, 45757280, 4321756, 43531566, 43531562, 43531588, 43531559, 45769906, 4221487, 37018912, 4130164, 45769875, 45770831, 45757445, 45757446, 43531608 |
| Type 2 diabetes drugs | 40164929, 40164897, 40163924, 1503297, 1525221, 19077638, 40164930, 19077636, 45774754, 45774442, 793152, 45774758, 40164898, 40163926, 45774893, 45774448, 19078552, 40169222, 19125043, 40169223, 40164946, 793147, 45774446, 45774438, 45774895, 1560171, 19125041, 1597772, 793143, 19079990, 19077682, 1597773, 37003617, 19080793, 1597761, 40163928, 19079293, 45892686, 19125049, 37003616, 19125051, 40239219, 19077637, 44785831, 37003607, 40170911, 19078551, 741834, 40220373, 45774435, 793154, 19078603, 19077659, 19006932, 1537597, 45774751, 1580747, 44785473, 780249, 40220376, 1559684, 40239218, 40166816, 19078530, 19030445, 43013924, 780260, 1525220, 1596916, 43526467, 46234237, 780256, 1597757, 44785477, 19009384, 46221581, 44785833, 1525216, 40167020, 40166814, 43526468, 1525215, 40220375, 19030443, 40167021, 1537599, 1525217, 19125045, 19125047, 1597756, 1583722, 1537596, 1597760, 37003609, 793153, 44785829, 37496842, 40164925, 40164947, 1596960, 1537601, 1559685, 40220371, 40164892, 40163929, 40166041, 40166815, 780266, 1537603, 37496746, 1597759, 741832, 37496844, 37496840, 43526472, 40164891, 19023424, 43013911, 45774533, 1596957, 779705, 19023425, 1537605, 1583729, 43526471, 1537598, 37003606, 19012270, 19023063, 45774531, 40163927, 1586345, 19079465, 780262, 1503328, 1537602, 1525218, 40166002, 40166037, 1547504, 780248, 1583730, 40163922, 1537600, 780259, 19125223, 19023426, 1547510, 40165997, 19006910, 1529352, 1583727, 1537604, 40163923, 37003608, 19027257, 1518148, 40239216, 40163925, 1503327, 780255, 37496838, 1529331, 793114, 42708171, 40164923, 19135405, 43526465, 19054444, 780264, 19054443, 42708078, 42708079, 40164932, 1596956, 19125221, 40166042, 1516766, 793111, 40164882, 37499746, 43013928, 793327, 42708167, 1516771, 19054445, 1547506, 1502830, 780265, 19135263, 40167017, 37496832, 1560168, 780258, 19047663, 19021312, 1516770, 42708170, 40164922, 40164900, 40166003, 19047612, 780261, 40164881, 44816273, 793331, 40165975, 40167016, 46287691, 42708087, 40165972, 43013884, 1718604, 19125219, 43013925, 19006933, 40165998, 793329, 42708166, 46287410, 1718711, 793321, 19126618, 1592795, 1560233, 1502829, 19099245, 1547505, 1502826, 1547508, 42903113, 40164943, 19081295, 40164915, 19047638, 1718599, 1559784, 1592794, 19040969, 42708175, 44816274, 19117945, 40166038, 1502827, 45892179, 19099207, 19023585, 19099208, 19071495, 40164904, 40164894, 40166035, 40164944, 44816282, 45775999, 780263, 1559782, 1594973, 46287408, 45892176, 40063354, 40165994, 793293, 42708086, 42708174, 19099209, 1547507, 45775621, 45774714, 19030580, 1592800 |

(continued)

**eTable 4. Condition-Related OMOP Concept IDs (continued)**

| Type 2 diabetes drugs (continued) | 19117200, 45775455, 42708089, 1592798, 19001408, 19099246, 43013912, 1592804, 1592803, 44816283, 42902992, 40165976, 43013896, 780257, 40164948, 45774721, 40164905, 45775860, 1718706, 40165973, 45892172, 19030579, 40167636, 1592653, 46287689, 1592656, 46234234, 19077684, 45774722, 19025946, 40164938, 19040970, 19077681, 46287684, 1515107, 45774710, 44816332, 40168374, 40229046, 19120648, 19030575, 19062999, 40229045, 1515249, 36249798, 19085004, 40164942, 45892177, 45774719, 40165995, 1596918, 42708088, 40164917, 42902823, 19117198, 1559760, 40167022, 40164889, 40164886, 42903382, 37497424, 19120315, 36249793, 19134639, 40229047, 19010532, 43013929, 19010204, 46287688, 43013899, 42708091, 43013897, 40171448, 41348912, 40165970, 40229048, 40165969, 19030572, 46287686, 40044251, 45774724, 37496783, 46287680, 40164913, 19078029, 19078030, 40063350, 45775867, 21110724, 19027259, 19121940, 793300, 42903059, 40164916, 45775623, 19012702, 40229050, 19078028, 19030576, 40164907, 19125042, 1597775, 793306, 19086484, 19045606, 40164888, 43013915, 37499835, 19012703, 21051793, 43013918, 793313, 19006908, 40164920, 19078607, 1718899, 1718901, 1502811, 43013900, 40164880, 45774717, 37496785, 40168370, 40168375, 1597792, 19060409, 19047829, 1510206, 40164941, 19029029, 19022851, 40164910, 40171449, 19047828, 1502809, 40166004, 21159907, 40166044, 43013916, 44057249, 19088818, 42708090, 1547554, 37499837, 1594976, 19029030, 1597776, 40164914, 45776000, 19045630, 19021786, 1525242, 1510207, 40164919, 19120257, 1718905, 40164885, 19079986, 40171451, 793828, 1718908, 40164918, 40220862, 19122368, 40164903, 36249791, 43013919, 45774715, 40167637, 40139022, 1515250, 19082950, 40099898, 793309, 19078604, 37499839, 1718904, 40231396, 40167012, 1597795, 19078553, 19045629, 1515254, 19125040, 1597774, 19024509, 1718912, 40168371, 1525244, 40164911, 793826, 19021787, 45775619, 40229049, 19117199, 19045625, 43013921, 40164924, 40046201, 19045626, 40167013, 19134624, 1515251, 19134642, 19135386, 19081296, 1510208, 19120166, 1718898, 19024508, 19099070, 1597798, 1529358, 19006909, 40168368, 43013905, 1525243, 19030573, 1597799, 40167634, 1597801, 19116559, 1596974, 40221118, 1592323, 19029061, 40239222, 42902866, 1502812, 43013902, 43011498, 21140107, 1596973, 19024503, 45775865, 45775622, 1502810, 40168369, 43013903, 40164890, 21149954, 42901620, 1529359, 1529360, 36249796, 42901622, 1596959, 43011499, 40170912, 1594975, 21169699, 1515258, 19082961, 19060491, 1718909, 1559785, 40059665, 44506754, 19122367, 46234240, 40165977, 19024507, 19024510, 1516794, 40231393, 1515253, 19099055, 1597781, 1559761, 1559728, 40164884 |
| --- | --- |

Abbreviations: OMOP, Observational and Medical Outcomes Partnership.

**eTable 5. Charlson Comorbidity Index Scoring System**

| **Condition number** | **Condition description** | **Points** |
| --- | --- | --- |
| 1 | Myocardial infarction | 1 |
| 2 | Congestive heart failure | 1 |
| 3 | Peripheral vascular disease | 1 |
| 4 | Cerebrovascular disease | 1 |
| 5 | Dementia | 1 |
| 6 | Chronic pulmonary disease | 1 |
| 7 | Rheumatic disease | 1 |
| 8 | Peptic ulcer | 1 |
| 9 | Liver disease, mild | 1 |
| 10 | Diabetes without chronic complications | 1 |
| 11 | Renal disease, mild to moderate | 1 |
| 12 | Diabetes with chronic complications | 2 |
| 13 | Hemiplegia or paraplegia | 2 |
| 14 | Any malignancy | 2 |
| 15 | Liver disease, moderate to severe | 3 |
| 16 | Renal disease, severe | 3 |
| 17 | HIV infection, no AIDS | 3 |
| 18 | Metastatic solid tumor | 6 |
| 19 | AIDS | 6 |

**eTable 6. Charlson Comorbidity Index Hierarchy Categories^2^**

| **Category** | **Hierarchy rule** |
| --- | --- |
| 1 | Hemiplegia/paraplegia (Condition 13) trumps cerebrovascular disease (Condition 4) |
| 2 | Liver disease, moderate to severe (Condition 5) trumps liver disease, mild (Condition 9) |
| 3 | Diabetes with complications (Condition 12) trumps diabetes without complications (Condition 10) |
| 4 | Renal disease, severe (Condition 16) trumps renal disease, mild to moderate (Condition 11) |
| 5 | Metastatic solid tumor (Condition 18) trumps any malignancy (Condition 14) |
| 6 | AIDS (Condition 19) trumps HIV (Condition 17) |

Note. Within each hierarchy, the milder condition does not contribute to the Charlson Comorbidity Index score if the more severe condition is present.

**eTable 7. ICD-9 and ICD-10 Codes for Charlson Comorbidity Index Conditions^2^**

| **Condition** | **ICD-9 diagnosis codes** | **ICD-10 diagnosis codes** |
| --- | --- | --- |
| Myocardial infarction | 410, 412, 410.x, 412.x, | I21, I22, I21.x, 122.x, 125.2 |
| Congestive heart failure | 398.91, 402.01, 402.11, 402.91, 404.01, 404.03, 404.11, 404.13, 404.91, 404.93, 425.4, 425.5, 425.6, 425.7, 425.8, 425.9, 428 | I11.0, I13.0, I13.2, I25.5, I42.0, I42.5, I42.6, I42.7, I42.8, I42.9, P29.0, I43, 150, I43.x, I50.x |
| Peripheral vascular disease | 093.0, 437.3, 443.1, 443.9, 447.1, 557.1, 557.9, V43.4, 440, 441, 440.x, 441.x, 443.2x, 443.8x | I73.1, I73.8, I73.9, I77.1, I79.0, I79.1, I79.8, K55.1, K55.8, K55.9, Z95.8, Z95.9, I70, I71, I70.x, I71.x |
| Cerebrovascular disease | 362.34, 430, 431, 432, 433, 434, 435, 436, 437, 438, 430.x, 431.x, 432.x, 433.x, 434.x, 435.x, 436.x, 437.x, 438.x | G45, G46, I60, I61, I62, I63, I64, I65, I66, I67, I68, G45.x, G46.x, G32.0x, H34.0x, H34.1x, H34.2x, I60.x, I61.x, I62.x, I63.x, I64.x, I65.x, I66.x, I67.x, I68.x |
| Dementia | 290.0, 290.3, 294.0, 294.8, 331.0, 331.2, 331.7, 797, 290.1x, 290.2x, 290.4x, 294.1x, 294.2x, 331.1x | F04, F05, F06.1, F06.8, G13.2, G13.8, G31.1, G31.2, G91.4, G94, R41.81, R54, F01, F02, F03, G30, F01.x, F02.x, F03.x, G30.x, G31.0x |
| Chronic pulmonary disease | 506.4, 508.1, 508.8, 490, 491, 492, 493, 494, 495, 496, 500, 501, 502, 503, 504, 505, 490.x, 491.x, 492.x, 493.x, 494.x, 495.x, 496.x, 500.x, 501.x, 502.x, 503.x, 504.x, 505.x | J68.4, J70.1, J70.3, J40, J41, J42, J43, J44, J45, J46, J47, J60, J61, J62, J63, J64, J65, J66, J67, J40.x, J41.x, J42.x, J43.x, J44.x, J45.x, J46.x, J47.x, J60.x, J61.x, J62.x, J63.x, J64.x, J65.x, J66.x, J67.x |
| Rheumatic disease | 446.5, 710.0, 710.1, 710.2, 710.3, 710.4, 714.0, 714.1, 714.2, 725, 714.8x, 725.x | M31.5, M35.1, M35.3, M36.0, M05, M06, M32, M33, M34, M05.x, M06.x, M32.x, M33.x, M34.x |
| Peptic ulcer | 531, 532, 533, 534, 531.x, 532.x, 533.x, 534.x | K25, K26, K27, K28, K25.x, K26.x, K27.x, K28.x |
| Liver disease, mild | 070.22, 070.23, 070.32, 070.33, 070.44, 070.54, 070.6, 070.9, 573.3, 573.4, 573.8, 573.9, V42.7, 570, 571, 570.x, 571.x | K70.0, K70.1, K70.2, K70.3, K70.9, K71.3, K71.4, K71.5, K71.7, K76.0,  K76.2, K76.3, K76.4, K76.8, K76.9, Z94.4, B18, K73, K74, B18.x, K73.x, K74.x |
| Diabetes without chronic complications | 250.8x, 250.9x, 249.0x, 249.1x, 249.2x, 249.3x, 249.9x | E08.0x, E08.1x, E08.6x, E08.8x, E08.9x, E09.0x, E09.1x, E09.6x, E09.8x, E09.9x, E10.0x, E10.1x, E10.6x, E10.8x, E10.9x, E11.0x, E11.1x, E11.6x, E11.8x, E11.9x,  E13.0x, E13.1x, E13.6x, E13.8x, E13.9x |
| Renal disease, mild to moderate | 403.00, 403.10, 403.90, 404.00, 404.01, 404.10, 404.11, 404.90, 404.91, 585.1, 585.2, 585.3, 585.4, 585.9, V42.0, 582, 583, 582.x, 583.x | I12.9, I13.0, I13.10, N18.1, N18.2, N18.3, N18.4, N18.9, Z94.0, N03, N05, N03.x, N05.x |

(continued)

**eTable 7. ICD-9 and ICD-10 Codes for Charlson Comorbidity Index Conditions^2^ (continued)**

| **Condition** | **ICD-9 diagnosis codes** | **ICD-10 diagnosis codes** |
| --- | --- | --- |
| Diabetes with chronic complications | 250.4x, 250.5x, 250.6x, 250.7x | E10.2x, E10.3x, E10.4x, E10.5x, E11.2x, E11.3x, E11.4x, E11.5x, E13.2x, E13.3x, E13.4x, E13.5x, E08.2x, E08.3x, E08.4x, E08.5x, E09.2x, E09.3x, E09.4x, E09.5x |
| Hemiplegia or paraplegia | 334.1, 342, 343, 344, 342.x, 343.x, 344.x | G04.1, G11.4, G80.0, G80.1, G80.2, G81, G82, G83, G81.x, G82.x, G83.x |
| Any malignancy | 180 – 195, 200 – 208, 199.1, 238.6, 180.x – 195.x, 200.x – 208.x, | C00 - C34, C37 – C41, C43, C45 – C58, C60 - C63, C76, C80.1, C81 - C85, C88, C90 - C99, C00.x - C34.x, C37.x - C41.x, C43.x, C45.x - C58.x, C60.x - C63.x, C76.x, C81.x - C85.x, C88.x, C90.x - C99.x, |
| Liver disease, moderate to severe | 456.0, 456.1, 572.2, 572.3, 572.4, 572.8, 456.2x | I86.4, K76.5, K76.6, K76.7, I85.0x, K70.4x, K71.1x, K72.1x, K72.9x |
| Renal disease, severe | 403.01, 403.11, 403.91, 404.02, 404.03, 404.12, 404.13, 404.92, 404.93, 585.5, 585.6, 588.0, V45.11, V45.12, V56.0, V56.1, V56.2, V56.31, V56.32, V56.8, 586, 586.x | I12.0, I13.11, I13.2, N18.5, N18.6, N25.0, Z99.2, N19, Z49, N19.x, Z49.x |
| HIV infection, no AIDS | 042 - 044 | B20-B22, B24 |
| Metastatic solid tumor | 196 – 198, 199.0, 196.x - 198.x | C77-C79, C80.0, C80.2, C77.x - C79.x |
| AIDS (HIV infection + opportunistic infection) | 117.5, 007.4, 078.5, 007.2, 136.3, V12.61, 046.3, 003.1, 799.4, 112, 180, 114, 348.3, 054, 115, 176, 031, 130, 200, 201, 202, 203, 204, 205, 206, 207, 208, 010 - 018, 112.x, 180.x, 114.x, 348.3.x, 054.x, 115.x, 176.x, 031.x, 130.x, 200.x, 201.x, 202.x, 203.x, 204.x, 205.x, 206.x, 207.x, 208.x, 010.x - 018.x | B37, C53, B38, B45, B25, G93.4, B39, C46, A31, B58, C81 - C96, A15, A16, A17, A18, A19, A07.2, B00, A07.3, B59, Z87.01, A81.2, A02.1, R64, B37.x, C53.x, B38.x, B45.x, B25.x, G93.4x, B39.x, C46.x, A31.x, B58.x, C81.x - C96.x, A15.x, A16.x, A17.x, A18.x, A19.x |

Abbreviations : ICD-9, International Classification of Diseases, 9th Revision; ICD-10, International Classification of Diseases, 10th Revision.

**eTable 8. Laboratory Tests and LOINC or SNOMED CT Codes**

| **Laboratory test** | **LOINC codes** | **SNOMED CT code** |
| --- | --- | --- |
| HbA1c | 4548-4, 17856-6 | 43396009 |
| HDL | 2085-9 | 28036006 |
| LDL | 13457-7, 2089-1, 18262-6 | 113079009 |
| Total cholesterol | 2093-3 | 121868005 |
| Triglycerides | 2571-8 | 14740000 |
| ALT | 1742-6, 1744-2, 1743-4 | 34608000 |
| AST | 1920-8 | 45896001 |
| Total bilirubin in blood | 1975-2 | n/a |
| Total bilirubin in urine | 5770-3, 50551-1 | n/a |
| GGT | 2324-2 | 69480007 |
| ALP | 6768-6 | n/a |
| CRP | 1988-5, 30522-7 | 55235003 |
| Red blood cell count (erythrocytes) | 789-8 | n/a |
| White blood cell count (leukocytes) | 6690-2 | n/a |
| Hemoglobin | 718-7 | n/a |
| Platelet count | 777-3 | n/a |
| Lipase | 3040-3 | 72680000 |
| Amylase | 1798-8 | 64435009 |
| Creatinine in blood | 2160-0, 38483-4 | n/a |
| Creatinine in urine | 2161-8 | n/a |
| eGFR | 77147-7, 48643-1, 48642-3, 98979-8, 69405-9 | n/a |
| Systolic blood pressure | 8480-6, 8459-0 | 271649006 |
| Diastolic blood pressure | 8462-4, 8453-3 | 271650006 |
| Heart rate | 8867-4 | n/a |
| Waist circumference | 56086-2 | n/a |
| Hip circumference | 62409-8 | n/a |

Abbreviations: ALP, alkaline phosphatase; ALT, alanine transaminase; AST, aspartate aminotransferase; CRP, C-reactive protein; eGFR, estimated glomerular filtration rate; GGT, gamma-glutamyl transferase; HbA1c, hemoglobin; HDL, high-density lipoprotein; LDL, low-density lipoprotein; LOINC, Logical Observation Identifiers Names and Codes; n/a, not applicable; SNOMED CT, Systemized Nomenclature of Medicine – Clinical Terms.

**eTable 9. Fitbit Wearable Sensor Data Analysis**

| **Fitbit Metrics** | **N** | **Pre GLP-1**  **initiation** | **Post GLP-1**  **initiation** | **Delta** | ***P* value** | **95% Confidence**  **interval** |
| --- | --- | --- | --- | --- | --- | --- |
| Daily minimum heart rate  (beats per minute) | 51 | 60.66 | 62.43 | 1.77 | 0.0027 | (0.64, 2.90) |
| Daily average heart rate  (beats per minute) | 51 | 80.78 | 81.78 | 1.00 | 0.0842 | (-0.14, 2.14) |
| Daily average step count (steps/day) | 79 | 7172 | 7163 | -9 | 0.9577 | (-339, 321) |
| Sleep (minutes/day) | 54 | 367.8 | 373.6 | 5.8 | 0.2560 | (-4.3, 15.8) |
| Restless sleep (minutes/day) | 54 | 14.7 | 13.7 | -1.0 | 0.3058 | (-2.78, 0.89) |

Abbreviations: GLP-1, glucagon-like peptide-1.
